# Deep learning applied to 4-electrode EEG resting-state data detects depression in an untrained external population

**DOI:** 10.1101/2022.03.28.22272733

**Authors:** Damian Jan, Manuel de Vega, Joana López-Pigüi, Iván Padrón

**Affiliations:** Instituto Universitario de Neurociencia, Universidad de La Laguna, 38200 La Laguna, Santa Cruz de Tenerife, Spain; Department of Psychology, Faculty of Health Sciences, University of Hull, Kingston upon Hull, United Kingdom

**Keywords:** Depression, Deep learning, Long short-term memory, Non-linear features, Diagnostic tool

## Abstract

In this study we trained and tested several deep learning algorithms to classify depressive individuals and controls based on their electroencephalography data. Traditionally, classification methods based on electroencephalography resting-state are based primarily on linear features or a combination of linear and non-linear features. Based on different theoretical grounds, some authors claim that the more electrodes, the more accurate the classifiers, while others consider that working on a selection of electrodes is a better approach□. In this study, a data-driven approach was initially applied on a selection of electrodes to classify 25 depressive and 24 control participants. Using a classifier with just four electrodes, based on non-linear features with high temporo-spatial complexity, proved accurate enough to classify depressive and control participants. After the classifier was internally trained and tested, it was applied to electroencephalography resting-state data of control and depressive individuals available from a public database, obtaining a classifier accuracy of 93% in the depressive and 100% in the control group. This validates the generalizability of the classifier to untrained data from different teams, populations and settings. We conclude that time-window span analysis is a promising approach to understand the neural dynamics of depression and to develop an independent biomarker.

## Introduction

Depression is a common mental disorder primarily characterized by a depressed mood and anhedonia, or loss of interest or pleasure. Other secondary symptoms such as appetite or weight changes, sleep difficulties (insomnia or hypersomnia), psychomotor agitation or retardation, fatigue or loss of energy, diminished ability to think or concentrate, feelings of worthlessness or excessive guilt and suicidality have also been reported [1]. Intensive research is being carried out on its possible causes (genetic, neurological, stress-related, inflammatory, immunological, microbiome-related, etc.), using a variety of methodological and disciplinary approaches (electroencephalography or EEG, neuroimaging, clinical medicine, psychiatry, biology, animal models, drugs, etc.). Yet, no conclusive unifying criteria or standardized biomarkers have been found [2].

It is estimated that 3.8% of the global population is affected by depression, including 5% of adults and 5.7% of adults over 60. Globally, 280 million people suffer from depression, which has been included as one of the critical/imperative conditions covered by WHO’s Mental Health Gap Action Programme (mhGAP) [3]. The Global Burden of Diseases, Injuries and Risk Factors Study showed that depression caused 34.1 million of the total years lived with disability (YLD), ranking as the fifth largest cause of YLD overall [4]. Furthermore, depression is a concomitant factor in different mental health conditions, like Adjustment Disorder [5], Alzheimer’s Disease [6], and others. With depression increasingly being a public health issue, its early detection is an important factor in reducing overloaded primary care systems and improving quality of life [7].

Depression is mainly diagnosed through questionnaires and clinical interviews based on the expertise of professionals with the guidance of WHO’s International Classification of Diseases (ICD 11) or the DSM-V in the USA. However, such an approach does not provide an entirely reliable diagnosis, since patients’ answers about their well-being or expectations of improving their mood are quite subjective, and also because emotional links between patients and doctors could be involved. All that can lead to a biased interpretation of questionnaires, making it necessary to advance in early depression detection through biomarkers that could help both in the diagnosis and in the evaluation of treatment [8, 9].

The quest for objective diagnosis has led to the development of EEG-based analysis protocols like Event Related Potentials, Band Power, Signal Features, Functional Connectivity, and Alpha Asymmetry, with average classificatory accuracies of 90% [10]. At the same time, the large amount of data provided by EEG calls for the use of fast, automatic machine learning algorithms.

The computer implementation of Machine Learning (ML) algorithms to automatically classify large amounts of data is derived from early proposals such as the brain learning mechanisms hypothesized by Hebb in 1949 [11], or the development of the perceptron by Rosemblatt in 1957 [12]. Since then, the number of algorithms and neural architectures has grown and their use has spread into almost every domain of daily life. Deep Learning (DL), as a new area of ML, combines multiple layers of the same or different basic architectural modules that have achieved goals such as speech recognition, translation or even the creation of online chatbots that help people to improve their mental health [13, 14]. The success of DL in these fields has encouraged many researchers to implement these methods for EEG data classification in the field of mental health [15].

A recent review on the use of resting-state EEG data classification for depression [16] pointed out the principal pitfalls and successes of research studies in this domain: 1) All used ML or statistical techniques for classifying, 2) Non-linear features worked best, 3) Electrode selection was based on theoretical criteria of the researchers and ranged from 1 to 30 electrodes, 4) Classification accuracy ranged from 80 to 99.5%, 5) None reported capacity of generalization of their classifiers to data from other sources, 6) The lack of public EEG databases with resting-state data on depressive individuals is an obstacle.

In our study, the ML and DL classifiers were initially applied to the resting-state EEG data obtained from a sample of university students with high and low scores in a depression questionnaire (BDI). We used a data-driven approach to electrode selection, to avoid a priori decisions about whether few or many electrodes are most appropriate. We selected only non-linear features, since this approach seems the most efficient strategy in depression research. Finally, we searched publicly available databases with EEG resting state to test whether the classifier trained with our participants’ data could be generalized to data from different populations.

## Methods

This study protocol was approved by the Human Research Ethics Committee of the University of La Laguna, Tenerife, Spain, with number CEIBA 2021-3100, to protect the participants’ rights according to the Declaration of Helsinki and was accepted by the board of the University Institute of Neuroscience (IUNE) of the University of La Laguna.

### Participants

A pre-screening was carried out with a sample of about 370 students of the University of La Laguna, who filled out an online version of the Beck Depression Inventory II, herein the BDI [17]. The BDI consists of 21 items and assesses the severity of symptoms in three categories using the following scoring: minimal depression 0-13 points, mild depression 14-19 points, moderate depression 20-28 points, and severe depression more than 29 points. We selected as depressive those participants who fell within the moderate and severe depression categories. The depressive group (DEP) included 26 participants (23/3 females/males; 22/4 right-/left-handed), with mean BDI scores of 32.92 (SD = 5.96) and a mean age of 22.43 years (SD = 1.4). The control group (CTL), with 24 participants (14/10 females/males; 23/1 right-/left-handed), had mean BDI scores of 3.87 (SD = 2.32) and a mean age of 19.6 years (SD = 2.5). The two groups showed statistical differences in BDI scores: *t*(48) = 22.32, p < .001, but did not differ in age: *t*(48) = 1.28, p > .05. All participants had normal or corrected-to-normal vision and hearing. The participants signed an informed consent form before the experiment and completed the Edinburgh Handedness Inventory [18].

### Procedure

#### EEG recording

EEG and EOG signals were recorded with an Easycap of 60 electrodes arranged according to the 10-20 standard system; 2 electrodes placed up and below the left eye used to keep control of blinks and vertical eye movements (EOG); and 2 additional electrodes placed on the left and right mastoids (M1/M2) as references. The EEG and EOG signals were amplified at 500Hz sampling rate using Synamp2 amplifier (Neuroscan; Compumedics), with high- and low-pass filters set at 0.05Hz and 500Hz, respectively. EEG electrode impedance was kept less than 10kΩ during the whole experiment.

#### Resting state

After the electrode set-up and impedance calibration, the participants were asked to stay calm and keep their eyes on a fixation point while we recorded EEG during 3 minutes for the Eyes Open Resting State condition (EO). Afterwards, they were asked to close their eyes and stay as calm as possible for 3 minutes while we recorded the EEG Eyes Closed Resting State condition (EC). For this analysis we used only time segments of the EC condition, as it seems to be more accepted for classifying depression [10].

#### Preprocessing

One participant of the DEP group was discarded due to corrupted data making it impossible to read the file. The total number of participants processed were 49 (25 DEP and 24 CTL).

The collected EEG data were preprocessed automatically for each participant with MNE-Python [19], discarding bad (noisy or flat) channels and noisy trials/epochs (noisy or with amplitude > 150 µV) and then eliminating blinks through the use of EOG channel (VEO) using the “mne.preprocessing.find_eog_events” function from the MNE package; no EEG channel was declared as miscellaneous (HEO, EKG, EMG, etc.). We used an average reference and a lower and higher passband from 0.1Hz to 200Hz. We applied a notch filtering of 50Hz and a low-pass band filtering to attenuate the frequencies above cut-off (90Hz). Once the whole EEG data had been preprocessed, they were epoched with a fixed time window that varied from 5s to 15s, following suggestions that this amount of time keeps data related to functional connectivity and network organization on the brain [20]. Finally, we kept a time window of 15s because it showed an increase in accuracy to the highest values in our study.

#### Feature extraction and selection

A group of 6 non-linear features, further explained below, were extracted with the “antropy” Python package, developed by Raphael Vallat [21]. The selected features were obtained from the 4 selected electrodes after applying the guidance from Giacometti [22]: antero-frontal 3 (AF3), frontal 5 (F5), antero-frontal 8 (AF8), and frontal 6 (F6) on epochs of 15s.

As a starting point, we used a logistic regression (LR) with a standard scalar (M = 0, SD = 1) as a baseline classifier, choosing the most relevant non-linear features (Higuchi’s fractal dimension and Sample Entropy) suggested by Čukić [16]. More features were subsequently added to improve classification. The study selected the following features for analysis:

- Higuchi’s Fractal Dimension (HFD) is an approximate value for the box-counting dimension used on fractal analysis to graph a real-valued time series. It has been used for more than 20 years in neurophysiological domains [23].
- Spectral Entropy (SPCTE) describes the complexity of a system based on applying the standard formula for entropy to the Power Spectral Density of EEG data. It quantifies the irregularity of EEG data [24].
- Permutation Entropy (PE) gives a quantification measure of the complexity of a dynamic system by capturing the order relations between values of a time series and extracting a probability distribution of the ordinal patterns [25].
- Singular Value Decomposition Entropy (SVDE) characterizes information content or regularity of a signal depending on the number of vectors attributed to the process [26].
- Sample Entropy (SE) is a modification of approximate entropy used for assessing the complexity of physiological time series signals [27].
- Detrended Fluctuation Analysis (DFA) is a stochastic process, chaos theory and time series analysis. DFA is a method for determining the statistical self-affinity of a signal. It is useful for analyzing time series that appear to be long short-term memory processes [28].

#### Testing classifiers

The data were normalized through Normalize scalar with an L2 metric. We applied a 10-fold cross validation test harness through StratifiedKfold from the Scikit-learn Python package [29].

A comparison analysis between ML (Logistic Regression - LR, Support Vector Machine - SVM) and DL algorithms (Multilayer Perceptron - MLP, Convolutional Neural Network - CNN, Long Short-term Memory - LSTM) was performed to check which approach gave the best classification scores. ML approaches were based on Scikit-learn Python packages while DL approaches were based on the Keras Python package [30]. LSTM obtained the highest classification scores (see Results).

#### Tuning LSTM

After tuning parameters, a stacked LSTM architecture with 60 training epochs and a batch size of 32 cross-entropy as loss function [31] and Adam optimizer [32] were obtained. A check pointer was used to save the best weights to reconstruct the model for predictions without the necessity of retraining it.

Data were EEG-epoched in segments of 15 seconds, normalized with L2 metric before calculation of features, split on 80% as training set and 20% as validation set. We found that considering the features as sequential “words” of one generic feature produced better outcomes in our analysis.

#### Validating with external data

Validating any classification approach with data provided by other researchers or that come from public repositories is a must for replication. To validate the present approach, we downloaded a database from PRED-CT, EEG Resting State Depression and Controls data [33]. The database used as external data were also divided in two groups; external Control group (n = 75, M = 1.73, SD = 1.65) and external Depressive group (n = 30, M = 25.10, SD = 3.19); a t-contrast test in BDI scores gave *t*(183) = −49.12, p < 0.001. We used as cut-off point a score of 25 in the Beck Inventory of Depression, that is, DEP ≥ 25 and CTL <= 7.

## Results

### Identifying key electrodes

In order to detect which group of electrodes was more accurate to classify controls/depressive participants, we applied an exploratory loop following Desikan’s brain structural areas mapped on associated electrodes as shown in Giacometti [22].

A 5-fold cross-validation logistic regression (LR) classification on three non-linear features (HFD, SE, SVDE) was systematically applied on the associated regions-electrodes as an exploratory baseline (see Table 1).

**Table 1.** Associated regions and electrodes as an exploratory baseline.

| Region | Electrodes | Cross-validation score |
| --- | --- | --- |
| Transverse Temporal | TP8, T7, T8 | 0.458 ($\pm 0.06$) |
| Banks Superior Temporal Sulcus | P8, TP7, TP8 | 0.415 ($\pm 0.1$) |
| Caudal Anterior Cingulate | FC2, AFz, F2 | 0.510 ($\pm 0.07$) |
| Caudal Medial Frontal | FC6, FC3, FC4 | 0.565 ($\pm 0.07$) |
| Isthmus-Cingulate | PO8, PO7, Pz, POz | 0.463 ($\pm 0.06$) |
| Lateral Occipital | PO7, PO8, O1, O2 | 0.481 ($\pm 0.04$) |
| Lateral Orbitofrontal | N2, F8, N1, F9, F10 | 0.601 ($\pm 0.08$) |
| Lingual Gyrus | Iz, PO8, PO7, Oz | 0.426 ($\pm 0.04$) |
| Medial Orbital Frontal | Fp2, N2, Fpz | 0.451 ($\pm 0.05$) |
| Paracentral Lobule | C1, C2, Cpz, Cz | 0.422 ($\pm 0.06$) |
| Pars Opercularis | FC5, FC6, FT7, FT8 | 0.544 (± 0.05) |
| Pars Orbitalis | F8, AF7, F9, F10 | 0.578 (± 0.05) |
| Pars Triangularis | FT7, FT8, F5, F8, F7 | 0.565 (± 0.08) |
| Pericalcarine | POz, O1, O2, Oz | 0.476 (± 0.06) |
| Postcentral Gyrus | T8, C5, C4, C6, C3 | 0.445 (± 0.03) |
| Posterior Cingulate | Cz, C1, FC2, C2 | 0.465 (± 0.06) |
| Precentral Gyrus | C3, C2, C4, C1 | 0.443 (± 0.07) |
| Precuneus | PO3, Pz, POz | 0.460 (± 0.07) |
| Rostral Anterior Cingulate | AF4, AFz, Fp2, Fpz | 0.526 (± 0.03) |
| **Rostral Medial Frontal Gyrus** | **AF3, F5, AF8, F6** | **0.658 (± 0.08)** |
| Superior Frontal Gyrus | AF3, F5, AF8, F6 | 0.494 (± 0.08) |
| Superior Parietal | POz, CP1, P2, P1 | 0.420 (± 0.04) |
| Superior Temporal Gyrus | TP8, FT9, T8, T7 | 0.458 (± 0.06) |
| Supramarginal Gyrus | C5, CP5, CP6 | 0.503 (± 0.08) |
| Cuneus | PO3, PO4, Oz, POz | 0.483 (± 0.09) |
| Inferior Parietal | P3, P4, P5, P6 | 0.481 (± 0.05) |

In bold, the region and the electrodes with the highest validation score.

Finally, electrodes associated with Rostral Medial Frontal Gyrus (AF3, F5, AF8, F6) were selected as the optimal region to continue the exploration for a good classifier.

### Comparing classifiers

We started the analysis with 6 selected non-linear features (HFD, SPCTE, SVDE, SE, PE, DFA) and applied a 10-fold cross validation test harness to ML and DL classifiers: Logistic Regression (LR), Support Vector Machine (SVM) with Radial Basis Function (RBF) Kernel, Multi-Layer Perceptron (MLP), Convolutional Neural Networks (CNN), and Long Short-term Memory (LSTM).

Performance of the classifiers was obtained by counting the hits, True Positives (TP) and True Negatives (TN), and the errors, False Positives (FP) and False Negatives (FN), and computing an accuracy index: Accuracy = (TP+TN) / (TP+TN+FP+FN). The results of the accuracy calculations were: LR = 0.763 (SD = 0.001); SVM = 0.813 (SD = 0.002); MLP = 0.837 (SD = 0.003); CNN = 0.821 (SD = 0.009); and LSTM = 0.916 (SD = 0.009). The independent t-tests with RStudio between performance of the classifiers confirmed that LSTM was the most efficient (see Fig 1). That is, accuracy was significantly larger for LSTM than for CNN (*t*(18) = −20.12, *p* < 0.001; Cohen’s *d* = 9); MLP (*t*(18) = 25.46, *p* < 0.001); Cohen’s *d* = 11.3); SVM (*t*(18) = 31.74. *p* < 0.001; Cohen’s *d =* 14.19); and LR (*t*(18) = −50.72438, *p* < 0.001; Cohen’s *d* = 22.68).

**Fig 1.**
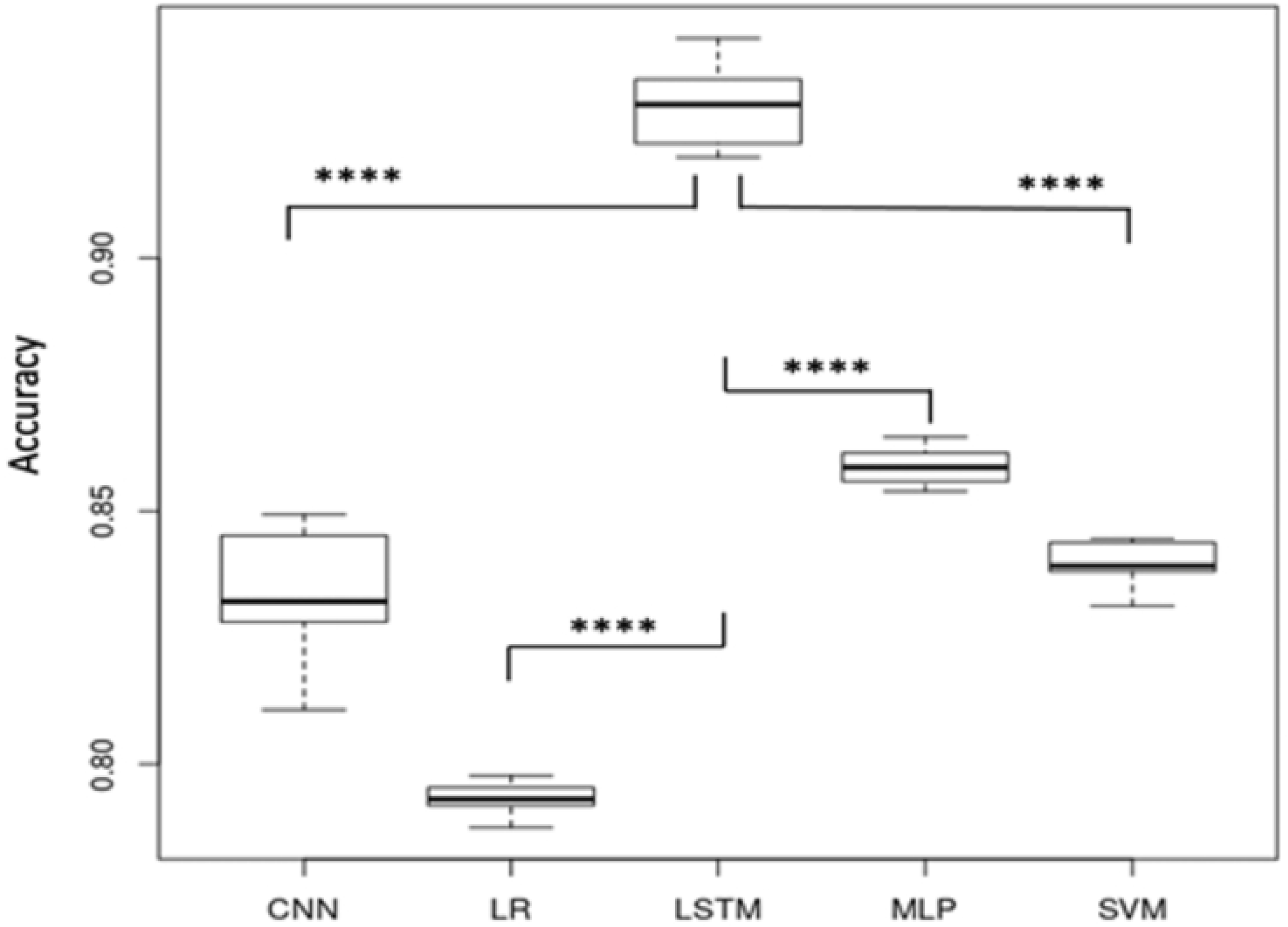
Accuracy comparison between the different classifiers. Convolutional Neural Networks (CNN), Logistic Regression (LR), Long Short-term Memory (LSTM), Multi-Layer Perceptron (MLP), Support Vector Machine (SVM). (**** *p* ≤ 0.0001)

### Tuned LSTM and validation with external data

Once LSTM had been selected as a target classifier, we explored its architecture and obtained a four-stacked LSTM architecture, that means 4 LSTM layers where the layer above provides a sequence output rather than a single value output to the LSTM layer below. Specifically, one output per input time step, rather than one output time step for all input time steps. A final Dense Layer eased the mapping over categories (CTL/DEP) (see Table 2).

**Table 2.** Four-stacked LSTM architectures with a final Dense Layer.

| Layer (type) | Output shape | Param# |
| --- | --- | --- |
| Lstm_20 (LSTM) | (None, 4, 100) | 40800 |
| Lstm_21 (LSTM) | (None, 4, 100) | 80400 |
| Lstm_22 (LSTM) | (None, 4, 100) | 80400 |
| Lstm_23 (LSTM) | (None, 100) | 80400 |
| Batch_normalization_2 (batch) | (None, 100) | 400 |
| Dense_3 (Dense) | (None, 1) | 101 |

Although the LSTM classifier outperformed with internal data, validation was needed before applying it to the external data. This was carried out through a process of trading off features (see Table 3) until an acceptable generalized model was found that could classify internal and public external data. The performance was categorized as follows. True Positive (TP): DEP people correctly identified as DEP; False positive (FP): CTL people incorrectly identified as DEP; True Negative (TN): CTL people correctly identified as CTL, and False Negative (FN): DEP people incorrectly identified as CTL. From these categories, some statistical variables were defined to choose the best combination.

**Table 3.** The process of validating the LSTM classifier with trading off features before applying it to external data.

| Feature | Accuracy | Precision | F1 | DEP ext. | CTL ext. |
| --- | --- | --- | --- | --- | --- |
| HFD, SE, DFA, PE, SVDE, SPCTE | 0.94 | 0.94 | 0.96 | 56% | 87% |
| HFD, DFA, SE | 0.76 | 0.72 | 0.77 | 37% | 93% |
| HFD, DFA, SVDE | 0.71 | 0.68 | 0.72 | 43% | 89% |
| HFD, SPCTE, SVDE | 0.77 | 0.75 | 0.77 | 50% | 62% |
| HFD, SE, PE, SVDE | 0.91 | 0.91 | 0.90 | 68% | 93% |
| HFD, SE, SVDE, SPCTE | 0.95 | 0.95 | 0.95 | 93% | 100% |

$* Recall / (Precision + Recall)$ combines precision and recall. Depressive group from external data (DEP ext.),
Control group from external data (CTL ext.).

Accuracy = (TP + TN) / (TP + TN + FP + FN)

Precision = TP/ (TP+FP) Recall = TP/(TP+FN)

F1 = 2 * Precision * Recall / (Precision + Recall), which combines precision and recall.

During such trade-off analysis we observed that DFA did not help in improving the results. Correlation analysis was applied to check possible distortions by such feature. Fig 2 shows that HFD and DFA were anticorrelative features. Lastly, it was decided to retain HFD, SE, SVDE, and SPCTE as final features for our model.

**Fig 2.**
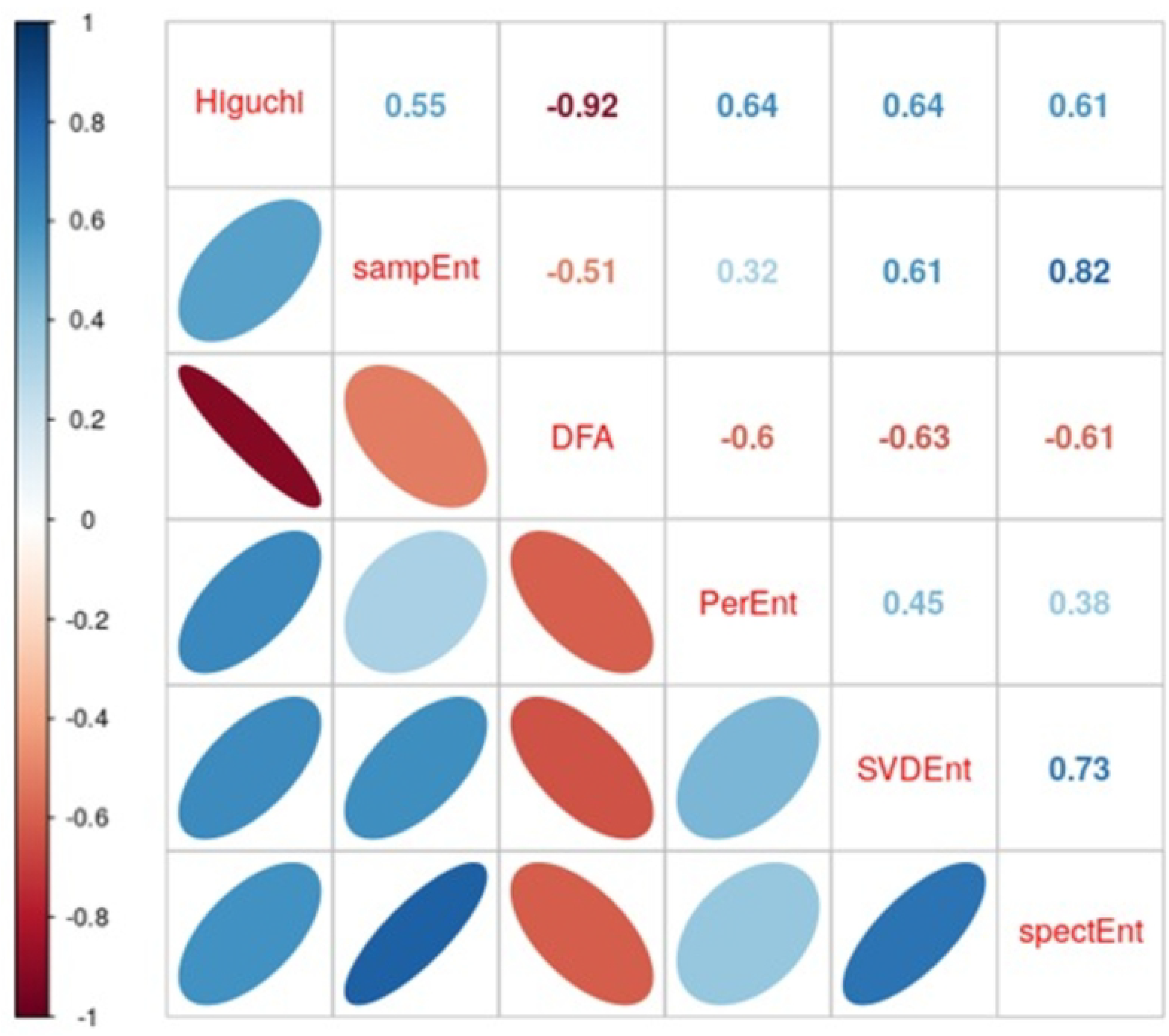
Correlation analysis between Higuchi Fractal Dimension (Higuchi), Sample Entropy (sampEnt), Detrended Fluctuation Analysis (DFA), Permutation Entropy (PerEnt), Singular Value Decomposition Entropy (SVDEnt), Spectral Entropy (SPCTE). Main diagonal represents features, correlation coefficients are shown in the upper triangular matrix, lower triangular matrix shows the graphic representation.

The graph of the loss function values as the training/validating progress during epochs (Fig 3) is used to detect overfitting and underfitting. In underfitting cases the validation curve is above the training one, while in overfitting, the validation curve, after matching the training in some point, starts to grow upward while the training curve remains low. In Fig 3, we see that both curves match each other so neither over- xnor underfitting affects the trained machine.

**Fig 3.**
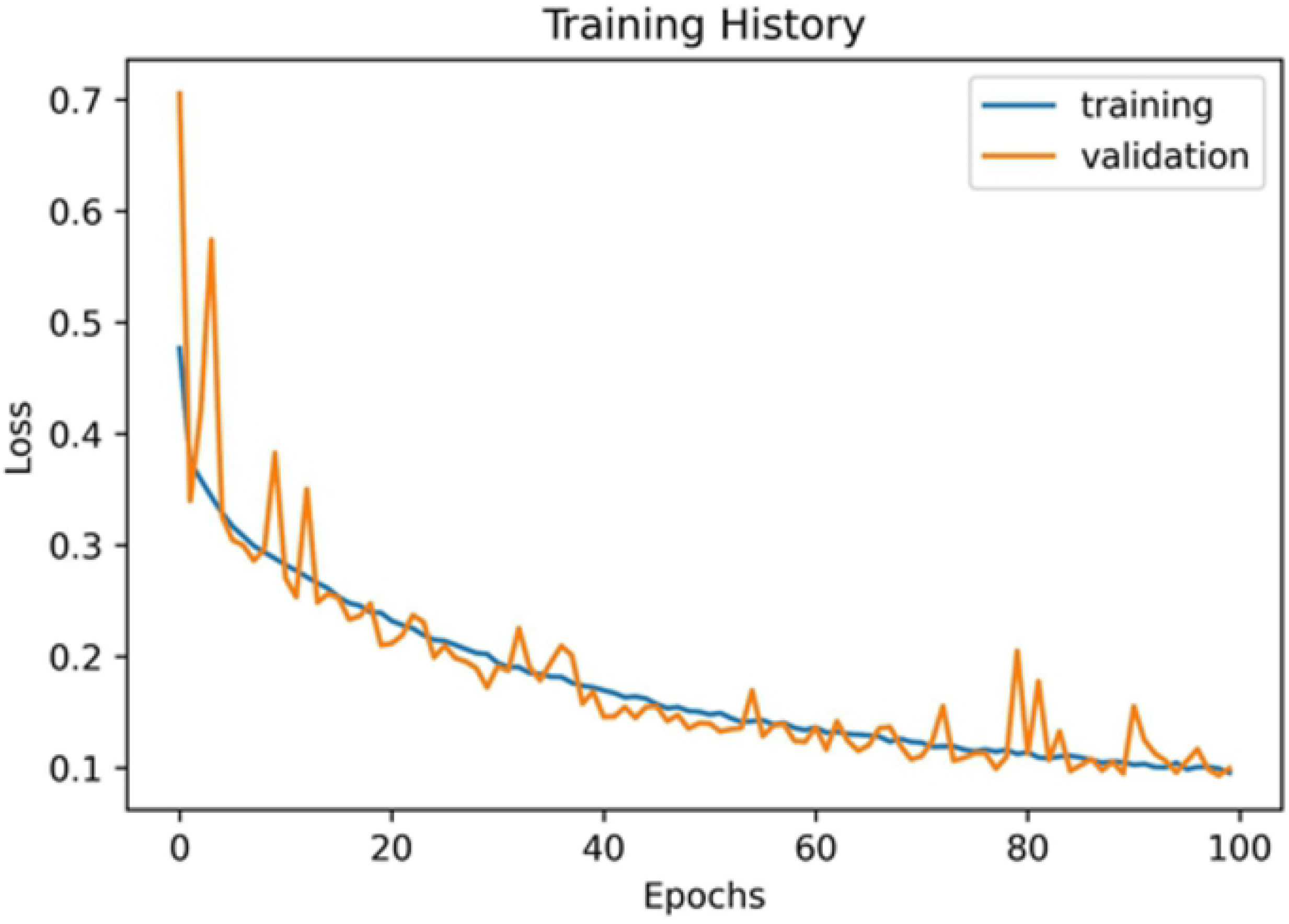
Training and validating history of the model with our sample of subjects. Loss is the value of the output function that minimizes the error between predicted and actual value.

Finally, applying the trained classifier to both local and external data we can obtain the Receiver Operating Characteristic (ROC) curves and the Area Under the Curve (AUC) to verify the goodness of the classifier [34]. Figs 4 and 5 show an AUC = 0.99 on local data and an AUC = 0.91 for external data. So the classifier can be considered a very good one.

**Figs 4 and 5.**
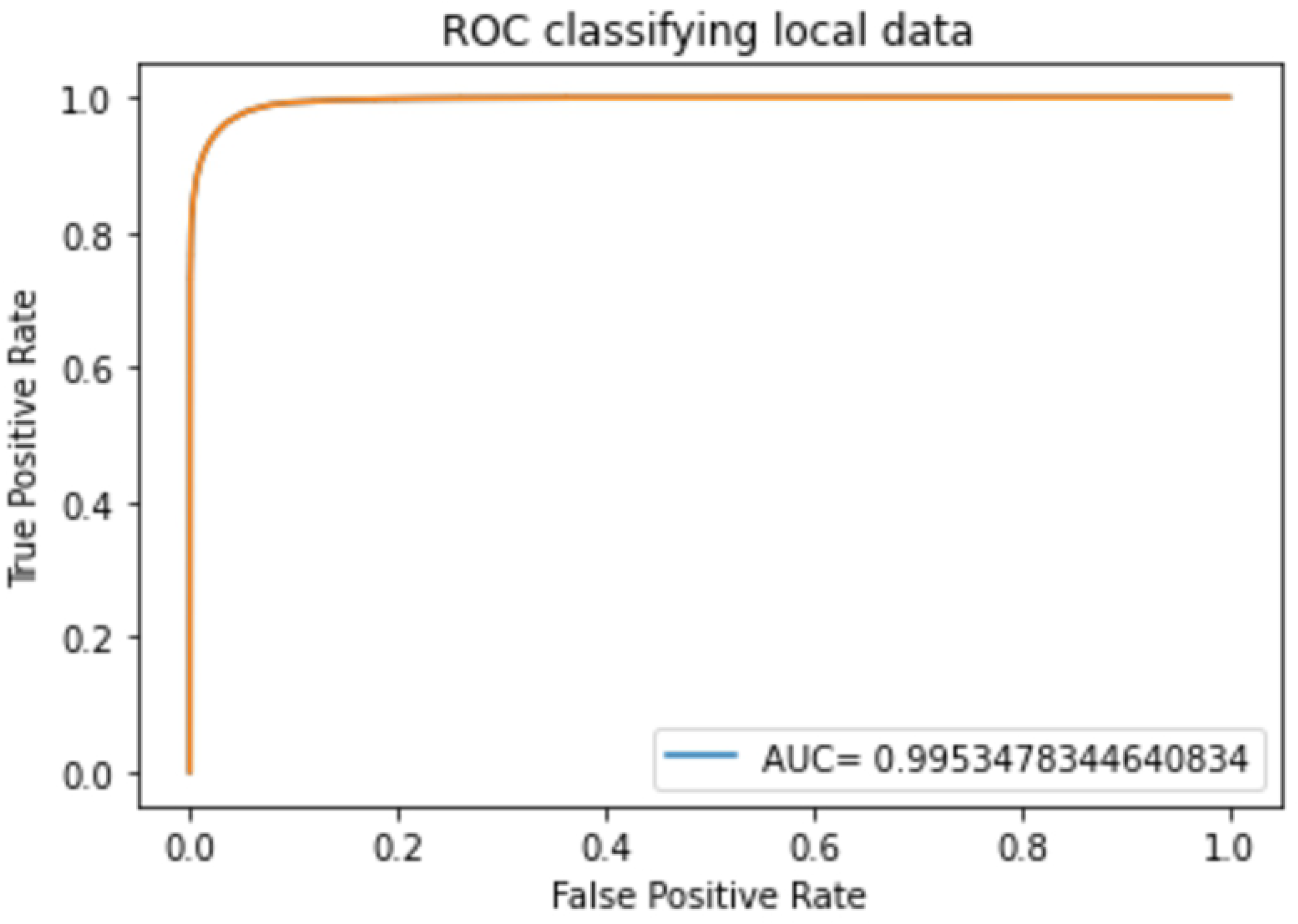
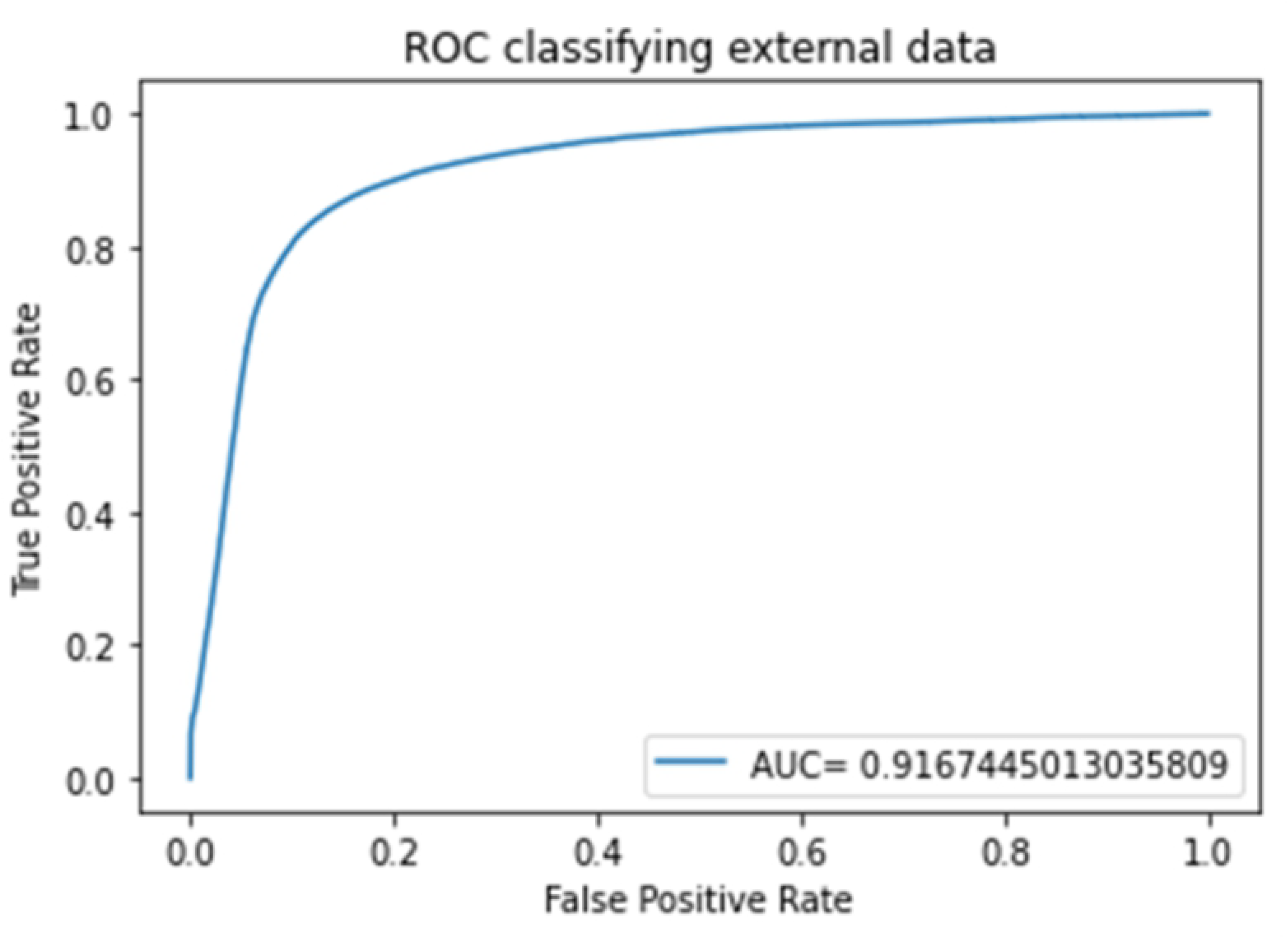
**Receiver Operating Characteristic (ROC) and Area Under the Curve (AUC) for classifying local data (above) and external data (below)**.

## Discussion

This study demonstrated that applying a deep learning-based classifier to EEG resting-state data, collected from a sample of college students, allows them to be accurately classified as depressed or non-depressed individuals. Furthermore, once the classifier was internally trained and tested with an accuracy of 95%, AUC = 0.99, it was successfully applied to detect depression in untrained EEG resting-state data from a publicly available source, with an accuracy of 93%, AUC = 0.91.

To achieve this remarkable performance, a bottom-up methodological approach, consisting of a selection of only four electrodes, and the combination of four non-linear features, was used to train and compare various machine learning and deep learning classifiers. It turned out that the LSTM model was the most accurate. This classifier keeps memory of the previous biosignal history, so it is especially appropriate in the analysis of time series where the hypotheses are related to time extended-signals, as is the case of the EEG resting state. Although our study was data driven, the fact that LSTM was the best classifier for depression suggested that the use of non-linear features in deep learning methods captures the long-standing and complex non-linear dynamics of the brain, serving as a potential diagnostic tool to detect abnormal EEG signatures associated with depression.

Our results show that a preselection of bands or linear features is not needed to efficiently classify depressive cases. Therefore, they do not support the hypothesis stated by Čukić et al. [16] that the more electrodes, the more information is retained to improve classification. On the contrary, we found that the proper choice of the time window epochs used to train the model is more crucial, as suggested by Fraschini et al. [20]. We can assume that the complexity displayed by the non-linear features is more related to changes in brain micro-states and more sensitive to time span than to location. This is the reason why techniques with high temporal resolution, such as EEG and MEG, are especially appropriate as diagnostic tools of brain dynamics in depression.

However, there is the possibility that the more structurally based a brain disease is, the more linear combined features need to be used to classify it. On the other hand, the more functionally based a disease is, the more non-linear combined features are necessary to classify it. From this point of view, we could think that depression seems to be a complex functional disease that cannot be accurately identified through simple linear features [16].

### Future research and limitations of the present study

This trained tool could be used to test improvements in depression diagnosis; a follow-up study using the trained classifier before and after treatment could be an interesting approach to test the tool as a guidance of objective treatment results, avoiding subjectivity in the evaluation process. This brings up the possibility to create a wearable, easy-to-use device that could be used in primary care settings to screen for depression, facilitating prevention.

Further research should be applied to distinguish among different types of depression, depression levels, and even anxiety and/or other comorbid disorders. Selective cognitive tasks could perhaps improve knowledge on depression if these new techniques are combined with traditional ones.

Moreover, further study of LSTM classification success should be analyzed to clarify the significance of time spanning signals.

The lack of more public databases with EEG resting state associated with depressive cases made it difficult to improve the classifier. Also, the present study was conducted on a young university population; to increase accuracy, a broader study with all ages is necessary.

## Conclusion

Our study demonstrates that using only non-linear features and few electrodes, we can obtain a good level of classification with generalized performance over external databases.

Deep learning approaches outperform machine learning ones, as they obtain better score classification values in this case (see Fig 1). Classifiers based on EEG data with few electrodes using Brain Computer Interfaces (BCI) are an easy tool to implement as support for diagnosis.

## Data Availability

All relevant data are within the manuscript and its Supporting Information files.

## References

1. American Psychiatric Association. Diagnostic and statistical manual of mental disorders. 2013.

2. Li Z, Ruan M, Chen J, Fang Y. Major Depressive Disorder: Advances in Neuroscience Research and Translational Applications. Neurosci Bull. 2021 Jun 13;37(6):863–80.

3. WHO. Scaling up care for mental, neurological, and substance use disorders. mhGAP Ment Heal Gap Action Program. 2008; Available from: http://www.who.int/mental_health/evidence/mhGAP/en/

4. James SL, Abate D, Abate KH, Abay SM, Abbafati C, Abbasi N, et al. Global, regional, and national incidence, prevalence, and years lived with disability for 354 diseases and injuries for 195 countries and territories, 1990–2017: a systematic analysis for the Global Burden of Disease Study 2017. Lancet. 2018 Nov 10 [Cited 2022 March 16]. Available from: https://linkinghub.elsevier.com/retrieve/pii/S0140673618322797

5. Fegan J, Doherty AM. Adjustment Disorder and Suicidal Behaviours Presenting in the General Medical Setting: A Systematic Review. Int J Environ Res Public Health. 2019 Aug 18;16(16):2967.

6. Cantón-Habas V, Rich-Ruiz M, Romero-Saldaña M, Carrera-González M del P. Depression as a Risk Factor for Dementia and Alzheimer’s Disease. Biomedicines. 2020 Oct 28;8(11):457.

7. Costantini L, Costanza A, Odone A, Aguglia A, Escelsior A, Serafini G, et al. A breakthrough in research on depression screening: From validation to efficacy studies. Acta Biomedica. 2021.

8. Merino Y, Adams L, Hall WJ. Implicit Bias and Mental Health Professionals: Priorities and Directions for Research. Psychiatr Serv [Internet]. 2018 Jun;69(6):723–5.

9. Janssens ACJW, Martens FK. Reflection on modern methods: Revisiting the area under the ROC Curve. Int J Epidemiol. 2020 Aug 1;49(4):1397–403.

10. de Aguiar Neto FS, Rosa JLG. Depression biomarkers using non-invasive EEG: A review. Neurosci Biobehav Rev. 2019 Oct;105:83–93.

11. Shaw GL. Donald Hebb: The Organization of Behavior. In: Brain Theory. Berlin, Heidelberg: Springer Berlin Heidelberg; 1986. pp. 231–3.

12. Keith D. Foote. A Brief History of Machine Learning. 2021.

13. Abd-alrazaq AA, Alajlani M, Alalwan AA, Bewick BM, Gardner P, Househ M. An overview of the features of chatbots in mental health: A scoping review. Int J Med Inform. 2019 Dec;132:103978.

14. Vaidyam AN, Wisniewski H, Halamka JD, Kashavan MS, Torous JB. Chatbots and Conversational Agents in Mental Health: A Review of the Psychiatric Landscape. Can J Psychiatry. 2019 Jul 21;64(7):456–64.

15. Craik A, He Y, Contreras-Vidal JL. Deep learning for electroencephalogram (EEG) classification tasks: a review. J Neural Eng. 2019 Jun 1;16(3):031001.

16. Čukić M, López V, Pavón J. Classification of Depression Through Resting-State Electroencephalogram as a Novel Practice in Psychiatry: Review. J Med Internet Res. 2020 Nov 3;22(11):e19548.

17. Beck, A. T., Steer, R. A., & Brown G. Beck Depression Inventory–II. 1996.

18. Oldfield RC. The assessment and analysis of handedness: The Edinburgh inventory. Neuropsychologia. 1971 Mar;9(1):97–113.

19. Gramfort A. MEG and EEG data analysis with MNE-Python. Front Neurosci. 2013;7.

20. Fraschini M, Demuru M, Crobe A, Marrosu F, Stam CJ, Hillebrand A. The effect of epoch length on estimated EEG functional connectivity and brain network organisation. J Neural Eng. 2016 Jun 1;13(3):036015.

21. Vallat R, Walker MP. An open-source, high-performance tool for automated sleep staging. Elife. 2021 Oct 14;10.

22. Giacometti P, Perdue KL, Diamond SG. Algorithm to find high density EEG scalp coordinates and analysis of their correspondence to structural and functional regions of the brain. J Neurosci Methods. 2014 May;229:84–96.

23. Kesić S, Spasić SZ. Application of Higuchi’s fractal dimension from basic to clinical neurophysiology: A review. Comput Methods Programs Biomed. 2016 Sep;133:55–70.

24. Inouye T, Shinosaki K, Sakamoto H, Toi S, Ukai S, Iyama A, et al. Quantification of EEG irregularity by use of the entropy of the power spectrum. Electroencephalogr Clin Neurophysiol. 1991 Sep;79(3):204–10.

25. Henry M, Judge G. Permutation Entropy and Information Recovery in Nonlinear Dynamic Economic Time Series. Econometrics. 2019 Mar 12;7(1):10.

26. Henry M, Judge G. Permutation Entropy and Information Recovery in Nonlinear Dynamic Economic Time Series. Econometrics. 2019 Mar 12;7(1):10.

27. Richman JS, Moorman JR. Physiological time-series analysis using approximate entropy and sample entropy. Am J Physiol Circ Physiol. 2000 Jun 1;278(6):H2039–49.

28. Zorick T, Mandelkern MA. Multifractal Detrended Fluctuation Analysis of Human EEG: Preliminary Investigation and Comparison with the Wavelet Transform Modulus Maxima Technique. Aegerter CM, editor. PLoS One. 2013 Jul 3;8(7):e68360.

29. Pedregosa F, Varoquaux G, Gramfort A, Michel V, Thirion B, Grisel O, et al. Scikit-learn: Machine Learning in Python [Internet].2012 Jan 2; Available from: http://arxiv.org/abs/1201.0490

30. Chollet F. Keras [Internet]. 2015. Available from: https://github.com/keras-team/keras

31. Bishop CM. Pattern Recognition and Machine Learning; 2006.

32. Kingma DP, Ba J. Adam: A Method for Stochastic Optimization. 2014 Dec 22.

33. Cavanagh JF, Napolitano A, Wu C, Mueen A. The Patient Repository for EEG Data + Computational Tools (PRED+CT). Front Neuroinform. 2017 Nov 21;11.

34. Janssens ACJW, Martens FK. Reflection on modern methods: Revisiting the area under the ROC Curve. Int J Epidemiol. 2020 Aug 1;49(4):1397–403.

